# Travel Time as a Predictor of Missed Appointments and Telemedicine Utilization in a Rural Outpatient Clinic: A Retrospective Cross-Sectional Observational Study

**DOI:** 10.64898/2026.03.20.26348551

**Authors:** Parker Graves, Cameron Jacobsen, Alan Ho, Dallin Johnson, Derek Weaver

## Abstract

**Background:** Rural populations face disproportionate barriers to healthcare access, often due to geographic isolation and limited provider availability. Prior studies have shown that increased travel time negatively affects appointment adherence. Telemedicine has emerged as a potential solution, but understanding its utilization in rural populations remains ongoing.

**Methods:** This retrospective cross-sectional observational study analyzed all scheduled appointments (n=5,548) from a single rural family medicine clinic in the Pacific Northwest United States during 2024. One-way travel times were calculated using the Google Maps Distance Matrix API and categorized into Short (<15 minutes), Medium (15-30 minutes), and Long (>30 minutes) commute groups. Proportions for utilization and cancellations of both telemedicine and in-person appointments were assessed across commute groups using chi-square tests (p < 0.05 considered significant).

**Results:** Overall, the proportion of cancellations were significantly higher among patients with Long commutes (36.2%) compared to Medium (31.0%) and Short (32.2%) commute groups (p < 0.001). Telemedicine utilization increased proportionately with commute time (7.7% for Long commute patients vs. 1.5% for Short; p < 0.001). However, telemedicine cancellation proportions did not significantly differ across groups (21.2% for Long, 13.3% for Medium, 17.0% for Short; p = 0.122), suggesting comparable telemedicine adherence regardless of distance. The proportions for in-person appointment utilization and cancellation were both greatest for the Short commute group.

**Conclusion:** Longer travel times are associated with increased appointment cancellations for rural patients, reinforcing travel burden as a key barrier to care. Telemedicine use increases with commute distance and demonstrates consistent adherence across groups, indicating its value as a tool to address rural healthcare gaps. These findings support the continued expansion of telehealth infrastructure to improve care for geographically isolated populations.

## Introduction

According to recent U.S. Census data, 20% of the population, roughly 60 million people or 1 in 5 Americans, reside in rural areas(1). Estimates have indicated that with only 9-10% of physicians practicing rurally, approximately 22 million rural residents are living in health professional shortage areas (HPSAs)(2). It is well documented that people living in rural communities are disadvantaged as it relates to receiving medical care. For example, it has been shown that rural residents have higher age-standardized mortality rates, maternal mortality rates and infant mortality rates than their urban counterparts(3-5). Other studies have also documented that rural communities have higher cancer prevalence and are more likely to be diagnosed at later stages(6-8). With the magnitude of this rural health gap, more research is warranted to better understand why this disparity exists and what can be done to overcome it. One way this has been done is by observing how travel times and appointment types affect the amount of cancellations for rural patients.

The current literature strongly supports a direct correlation between driving distance and the amount of cancellations in both rural and urban settings. A retrospective analysis of an urban pediatric clinic showed that appointment non-arrival was associated with both longer transportation times and lower socioeconomic status(9). Another study showed that patients travelling to their prenatal appointments were 5% more likely to show up late or 3% more likely to cancel for every 10 minutes of increased travel time(10). In rural settings, longer driving times have been associated with both reduced patient attendance and reduced health utilization, including less screening and rehabilitation appointments(11-13). Among the efforts made to reduce the negative impact of lengthy commute times, telemedicine has been shown to be a positive countermeasure.

In a rural population, one study suggested that telemedicine appointments increased access to care for rural residents as well as other hard-to-reach patients such as those with difficult work schedules, complex health challenges and those from areas of higher poverty or lower education(14). A narrative review of 15 articles also concluded that telemedicine resulted in positive outcomes for patients and healthcare providers including reduced travel times, reduced staffing costs, and additional provision of specialty help in areas where no specialists were available(15). Although telemedicine is proving to be an innovative way to overcome some barriers to healthcare, it is not without its own challenges, especially for rural populations. Additional obstacles to telemedicine generally include poor connectivity, limitations to physician training, diminished diagnostic capabilities, and impediments to the fiduciary relationship between doctor and patient(16). With the current disparities in rural healthcare and the challenges of telemedicine, more research is warranted to understand the existing situation and to assist in offering additional strategies.

This study aimed to evaluate the relationship between patient commute times as a predictor for appointment adherence in a rural primary care clinic. Additionally, the study aimed to assess how telemedicine has been utilized and if it assists in overcoming any differences in appointment attendance for patients who have to commute further for this population. This was accomplished by separating patients into three commute time groups and comparing the proportion of cancellations, as well as observing differences in telemedicine and in-person appointment utilization and cancellation for each group. This paper hypothesized that longer commute times would correlate with higher proportions of total cancellations and higher proportions of telemedicine utilization. In observing the utilization of telemedicine, the study hypothesized that telemedicine cancellations would be insignificant between commute groups. To assist in the study’s evaluation, descriptive data was gathered on the patient population and the varying types of cancellations. With this analysis, this study intends to add to the current literature that defines the rural health gap, as well as provide a heightened context in which providers and patients are striving to overcome the disparities that they face.

## Methods

### Study Design and Setting

This was a retrospective cross-sectional observational study conducted at a single rural family medicine clinic in the Pacific Northwest area of the United States. RUCA codes have been used to distinguish between metropolitan, urban and rural settings.(17) RUCA codes from 1-3 are described as metropolitan or urban while RUCA codes from 4-10 are considered rural. The clinic resides in a community with a RUCA code of 4, which is defined as a micropolitan area and within rural classifications. The clinic also has a significant rural catchment area with patients coming from additional communities that have RUCA codes between 4 and 10. Data were collected from all scheduled patient appointments within the 2024 calendar year.

### Data Collection

Appointment-level data were extracted from the clinic’s electronic health records (EHR), including patient demographics (age, sex, race/ethnicity, address information), appointment type, and cancellation status (including reason, if applicable). Telemedicine visits and in-person appointments were segregated into two comparison groups. Each patient’s address was divided into four fields: street address, city, state, and ZIP code. Patient data was de-identified by using assigned numerical IDs to be in compliance with the HIPAA privacy rules(18). After patient data were de-identified, these components were programmatically concatenated using Python to generate a complete address for each patient.

### Travel Distance and Time Calculation

To calculate the one-way driving distance and estimated travel time between each patient’s home and the clinic, the Google Maps Distance Matrix API (GMAPI) was used, a tool that provides real-time routing information based on the current road network. GMAPI has shown to be a robust tool in determining accurate transportation time and distance(19). The GMAPI provided the estimated driving distance (in miles) and travel time (in minutes) for each patient ID. If the GMAPI failed to return a valid result for an address (e.g., due to invalid formatting or missing data), the corresponding entry was flagged and excluded from analysis. Mean, standard deviation, and range were then calculated for travel times.

Travel time was then stratified into three groups:

1. Short Commute (SC): < 15 minutes
2. Medium Commute (MC): 15-30 minutes
3. Long commute (LC): > 30 minutes

### Statistical Analysis

All analyses in this section were conducted using Python (version 3.12.7) with *NumPy, SciPy* and *pandas* packages. Once patients were stratified into the three cohorts, cancellation proportions were computed for each commute group and appointment type. Associations between categorical variables were evaluated by performing a series of chi-square tests of independence. Specifically, the study tested for associations between commute group and overall cancellations, distribution of cancellation types, telemedicine and in-person appointment usage, and telemedicine and in-person cancellations. Subgroup analyses were conducted to compare cancellation proportions between pairs of commute groups (e.g., Short vs. Long). The association between appointment type and cancellation proportion was also assessed using a chi-square test across all appointment categories. For all tests, a p-value of <0.05 was considered statistically significant. Confidence intervals (95%) for proportions were calculated using standard error estimates derived from binomial variance.

## Results

### Patient Demographics

A total of 5,548 patients were included in the dataset, with a mean age of 29.0 years. Of these, 52.6% were female (n=2,920) and 47.4% were male (n=2,628). The majority of patients identified as “White” (78.5%, n=4353), followed by “Other Race” (14.9%, n=828), “Patient Declined” (3.9%, n=214), “Black or African American” (1.1%, n=59), “American Indian or Alaska Native” (1.0%, n=54), “Asian” (0.5%, n=28), and “Native Hawaiian or Pacific Islander” (0.2%, n=12). Patient demographics are listed in table 1.

**Table 1:**
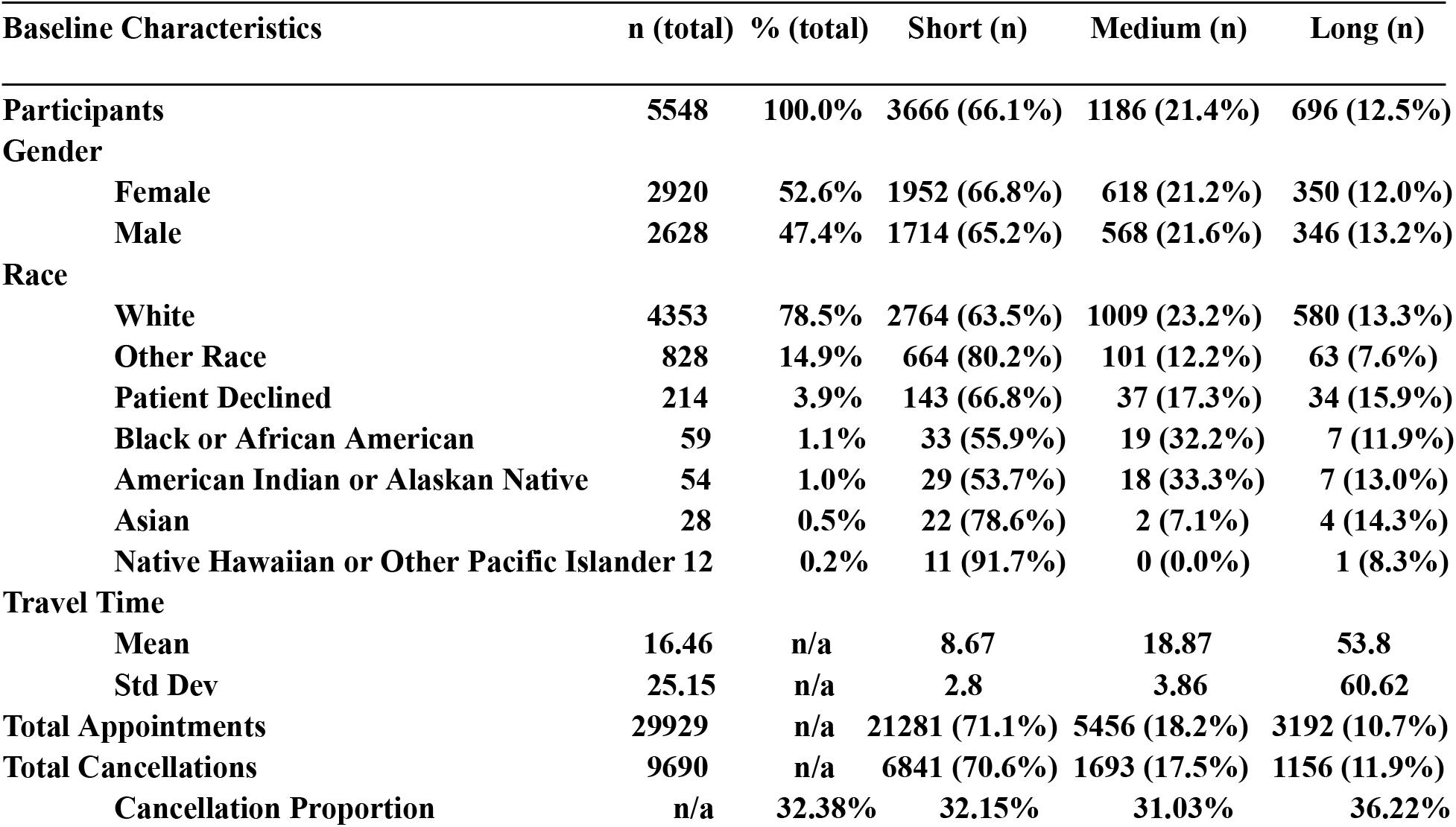
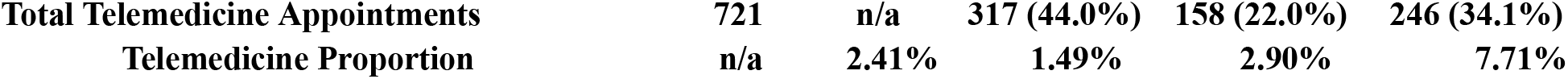
Population Demographics. Summary of patient demographics across all appointments scheduled during the study period. Values are reported as counts and percentages of the total study population (n = 5,548), as well as for each respective commute group.

### Total Cancellation Proportions and Types

Cancellation proportion varied by commute group, with patients in the Long commute group exhibiting the highest overall cancellations at 36.2% (± 0.9), followed by the Short commute group at 32.2% (± 0.3) and the Medium commute group at 31.0% (± 0.6). Chi-square testing revealed that the difference in cancellations across the three groups was statistically significant (p < 0.001), with the Long commute group having a significantly higher proportion of cancellations than both the Short and Medium commute groups. Figure 1 displays this trend, highlighting the positive correlation between increased travel time and likelihood of cancellation.

**Figure 1:**
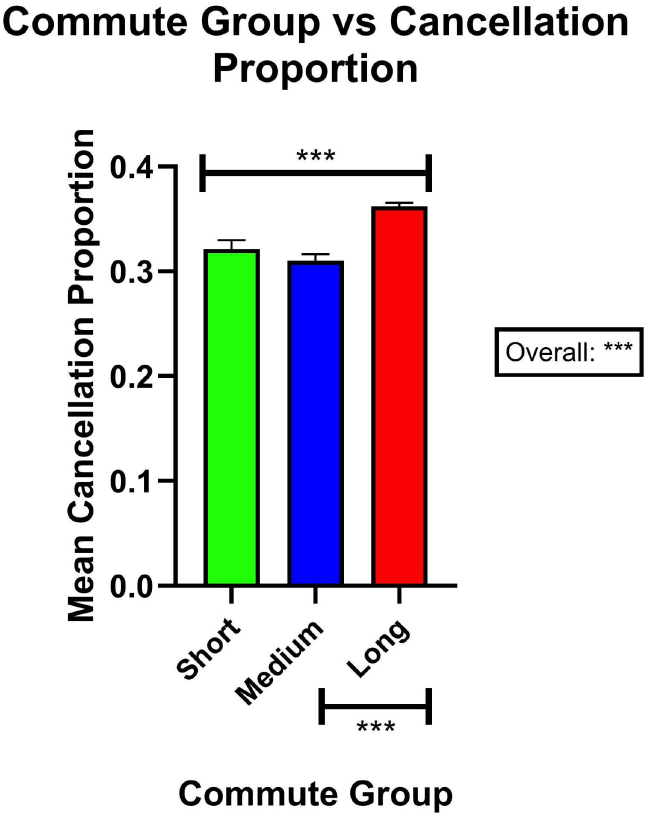
Commute Group vs Cancellation Proportion. This figure plots the mean cancellation proportion and SE for Short, Medium and Long commute groups. The mean cancellation proportions for each group is 0.322 ± 0.003, 0.310 ± 0.006 and 0.362 ± 0.009, respectively. Chi-square testing revealed that the distribution between the three groups is significantly different (p<0.001). Furthermore, additional Chi-square shows that the Long commute group had significantly higher cancellation proportions than both Short and Medium (p<0.001).

Additionally, the types of cancellations varied across commute groups, as shown in Figure 2. The distribution shows that commute groups had an effect on the reason for cancellation. The Short commute group had the highest percentage of no-show cancellations (35.0% ± 0.6), while the Medium commute group had the highest percentage of patient-initiated rescheduling (42.6% ± 1.2). In contrast, the Long commute group had the highest percentage of cancellations due to provider unavailability (8.5% ± 0.8) and other reasons (19.7% ± 1.2).

**Figure 2:**
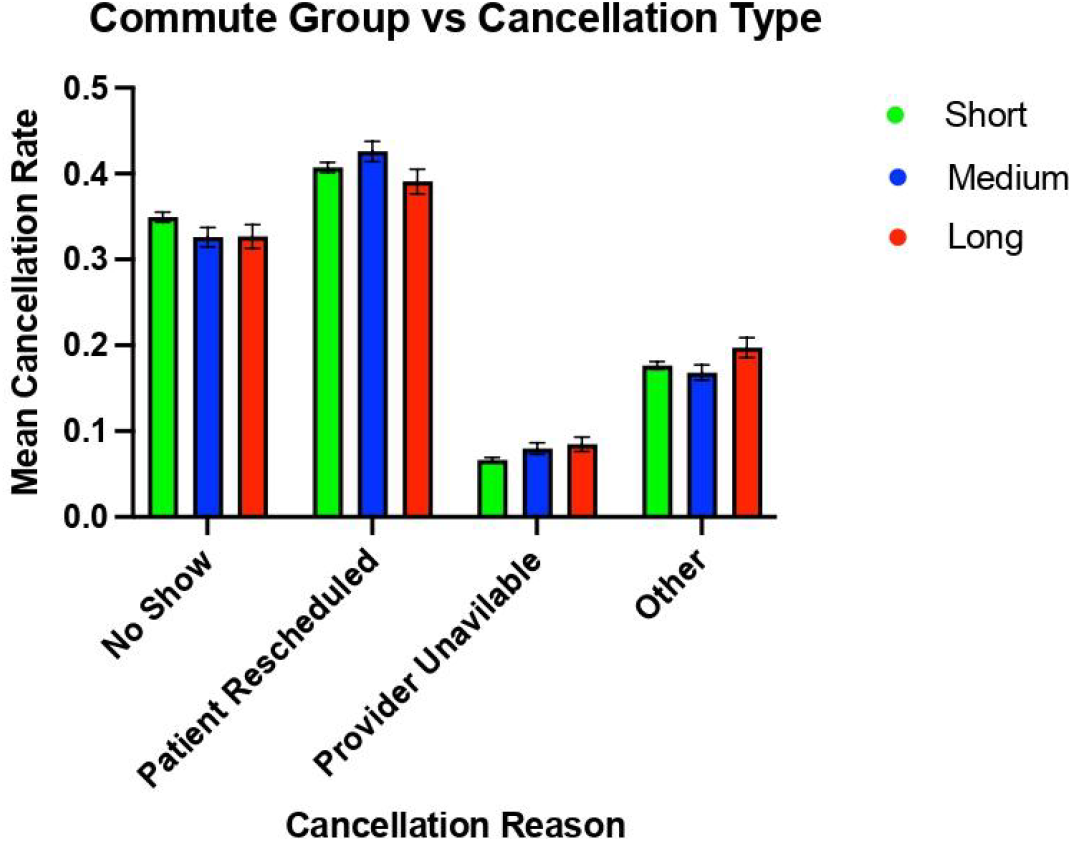
Commute Group vs Types of Cancellation. This figure plots the mean cancellation proportion and SE for each type of cancellation across each commute group. This distribution shows that commute groups had an effect on the reason for cancellation. For No Show cancellations, the Short commute group had the highest mean cancellation at 0.350 ± 0.006. For cancellations due to Patient Rescheduling, the Medium commute group had the highest cancellation proportion at 0.426 ± 0.012. For Provider Unavailability and Other cancellation reasons, the Long commute group had the highest proportion of cancellations at 0.085 ± 0.008 and 0.197 ± 0.012, respectively.

### Telemedicine vs In-Person Utilization and Cancellation Percentages

The percentage of telemedicine appointments relative to total scheduled visits increased significantly with commute time. Patients in the Long commute group utilized telemedicine at the highest percentage (7.7% ± 0.47), followed by the Medium (2.9% ± 0.23) and Short (1.5% ± 0.08) commute groups. As shown in Figure 3, these differences were statistically significant across all comparisons (p < 0.001), indicating a strong relationship between travel time and telemedicine utilization. Inversely, the percentage of in-person appointments decreased significantly as commute times enlarged. Patients in the Long commute group utilized in-person appointments at the lowest proportion (*92*.*3% ± 0*.*5*), with the Medium (*97*.*1% ± 0*.*2)* group showing increased usage and the Short (*98*.*5% ± 0*.*1*) commute group having the greatest proportion of in-person visits. As shown in Figure 4, these differences were significant across all comparisons (*p<0*.*001*), indicating an equally strong relationship between travel time and in-person appointment usage. These trends support the idea that patients facing longer commutes may prefer or rely more heavily on telemedicine as a healthcare delivery method while patients with shorter commute times utilize in-person appointments with greater frequency.

**Figure 3:**
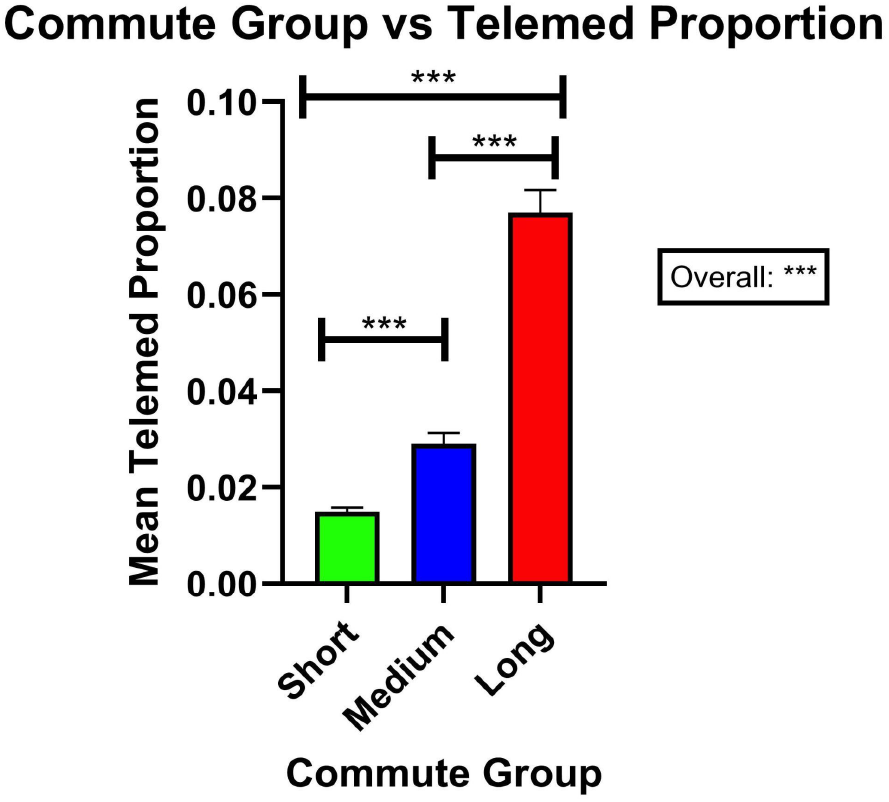
Commute Group vs Proportion of Telemedicine. This figure plots the proportion of telemedicine appointments out of all appointment types and SE for each commute group. Chi-squared testing showed that the distribution of telemedicine appointments was significantly impacted by commute group (p<0.001). In particular, as commute time increased, so did the proportion of telemedicine. The Long commute group had the highest usage of telemedicine at 0.077 ± 0.005 which was significantly higher than both Short and Medium commute groups (p<0.001). Furthermore, the Medium group’s proportion of telemedicine use at 0.029 ± 0.002 was also significantly higher than the Short group’s at 0.015 ± 0.001 (p<0.001).

**Figure 4:**
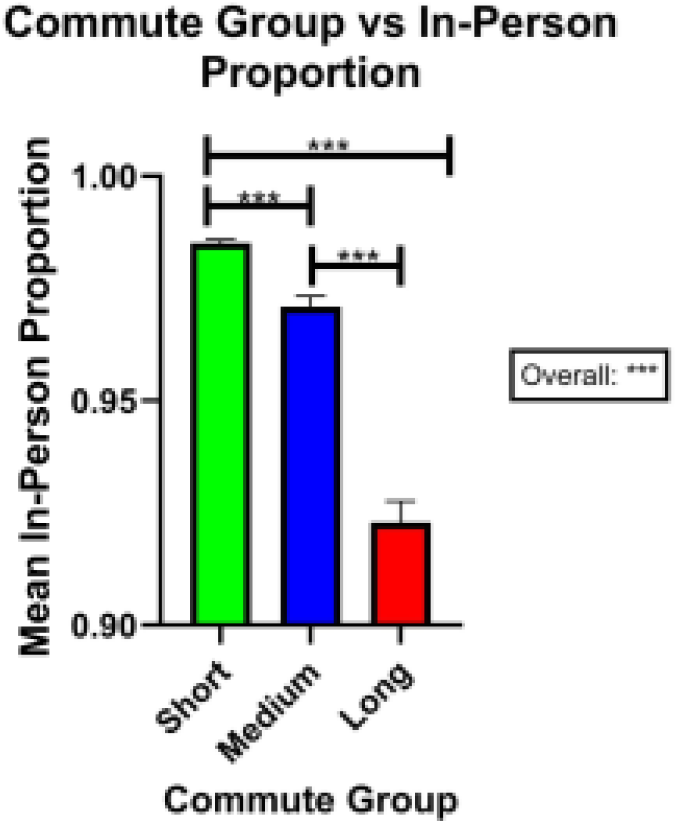
Commute Group vs Proportion of In-Person Appointments. This figure plots the proportion of in-person appointments out of all appointment types and SE for each commute group. Chi-squared testing showed that the distribution of in-person appointments was significantly impacted by commute group (p<0.001). In particular, as commute time increased, the proportion of in-person appointments decreased. The Long commute group had the lowest proportion of in-person appointments at 0.923 ± 0.005 which was significantly lower than both Short and Medium commute groups (p<0.001). Furthermore, the Medium group’s in-person use at 0.971 ± 0.002 was also significantly lower than the Short group’s at 0.985 ± 0.001 (p<0.001).

Despite the differences in telemedicine usage, the percentage of telemedicine cancellations did not differ significantly between commute groups. Figure 5 illustrates that the Long commute group had the highest telemedicine cancellation percentage at 21.2% (± 2.6), followed by the Short (17.0% ± 2.1) and Medium (13.3% ± 2.7) commute groups, though these differences did not reach statistical significance (p = 0.122). These findings suggest that while telemedicine is more frequently utilized by patients living farther from the clinic, their adherence to scheduled telehealth visits remains consistent with those in closer proximity, reinforcing telemedicine’s potential as a way to enhance rural healthcare delivery.

**Figure 5:**
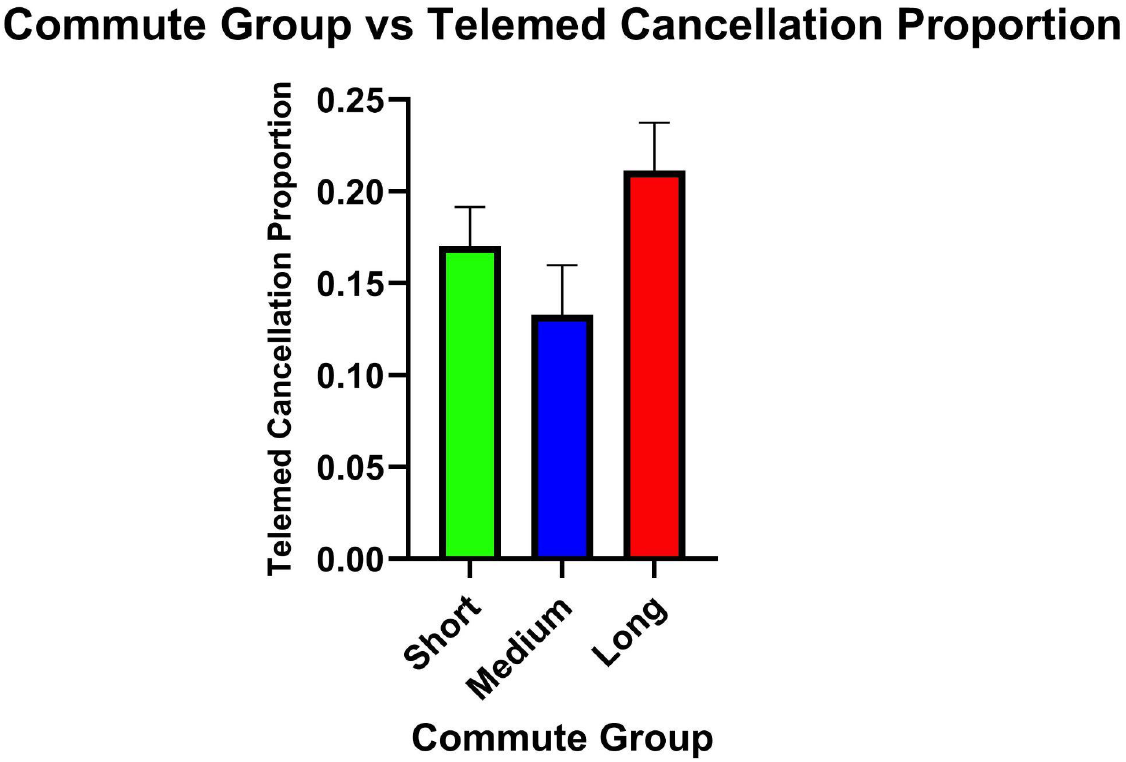
Commute Group vs Telemedicine Cancellation Proportion. This figure plots the proportion of telemedicine cancellations and SE across the three commute groups. The Long commute group had the higher proportion of telemedicine cancellations at 0.212 ± 0.026, followed by Short at 0.170 ± 0.021 and Medium at 0.133 ± 0.027. Chi-squared testing revealed no significant differences across the whole distribution or between commute groups (p=0.122).

Regarding in-person cancellations, Figure 6 shows that the Short commute group had a significantly higher percentage of cancellations than both the Medium (p<0.001) and Long (p<0.001) commute groups. The percentage of in-person cancellations for the Short commute group was 37.5% ± 0.9, followed by the Long (32.4% ± 0.3) and Medium (31.6% ± 0.6) commute groups, respectively. Together, these differences offer an interesting perspective on how commute time affects both telemedicine and in-person appointments in terms of utilization and cancellation.

**Figure 6:**
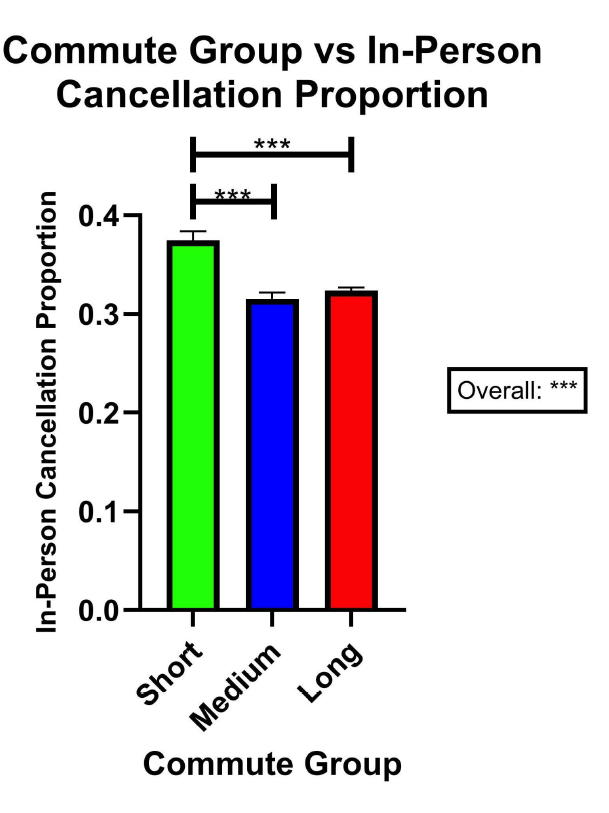
Commute Group vs In-Person Cancellation Proportion. This figure plots the proportion of in-person cancellations and SE across the three commute groups. The distribution of in-person cancellation is significantly impacted by the commute group (p<0.001). The Short commute group had the highest proportion of in-person cancellations at 0.375 ± 0.009, which was shown to be significantly higher than Medium (0.316 ± 0.006) and Long (0.324 ± 0.003) by Chi-Square testing (p<0.001). There was no significant difference between Medium and Long commute groups.

## Discussion

### Travel Times as a Predictor of Cancellations

The commute time of patients demonstrated to be a strong predictor for both the proportion and type of cancellations seen in the clinic. For the proportion of cancellations, the Short and Medium commute groups canceled at percentages of 32.1% and 31.0%, respectively, while the Long commute group canceled 36.2% of the time. This finding is supported by the current literature in that driving time is inversely proportional to appointment attendance. It’s important to note that within the literature, driving time, not necessarily driving distance, seems to correlate with greater cancellations and that this relationship is both multifactorial and not universally generalizable. As previously mentioned, Maldonado et al. (2020) showed that increased travel times correlated with a greater likelihood of patients being late or cancelling prenatal care visits in a rural area of North Carolina(10). Additionally, at an urban pediatric clinic, Wallace et al. (2018) displayed that patients with lower socioeconomic status and greater transit times had the largest rate of non-arrival, even when observing different travel modalities such as driving, public transit or walking(9). Both studies agree with this study’s findings concerning travel time and the proportion of cancellations regardless of whether they took place in a rural or urban setting. Paradoxically, one study that took place in California, being the most urban state according to US census data(20), found that a 15 min increase in travel time was associated with reduced risk of under or delayed treatment for early stage lung cancer patients(21). The different effects of travel times and appointment status in rural and urban areas suggest that there may be additional societal or structural factors at play. Nevertheless, this study, along with other rural studies, show that travel time continues to be a significant barrier to patients seeking medical care within rural communities.

Regarding the type of cancellations observed, there were significant differences between commute groups that cannot be fully explained from the data gathered. Those in the Short commute group were more likely to no show without notifying the clinic, patients in the Medium commute group were more likely to reschedule their appointment, and those in the Long commute group were more likely to notify the clinic to cancel their appointment. The significant differences between groups, though numerically small, suggests that how patients cancel their appointments is influenced by driving time. Although interesting, with the limited data collected this study is unable to extrapolate other potential reasons for why these differences may have occurred.

### Telemedicine and In-Person Appointment Utilization and Cancellation

In concordance with existing data, the study found that the Long commute group utilized telemedicine to a significantly larger proportion than either of the other commute groups. This was also reflected by the Long commute group utilizing in-person appointments significantly less than the Small and Medium commute groups. More specifically, the Long commute group was 2.7 times more likely to utilize telemedicine than the Medium commute group and 5.1 times more likely than the Short commute group. These findings are supported by multiple studies observing travel time and telemedicine utilization. For example, a report studying Federally Qualified Health Centers (FQHCs) demonstrated a dose-response correlation in regards to driving distance and telemedicine usage(22). Another study which utilized data from the Veterans Health Administration (VA) found that telemedicine encounters significantly increased patients’ access to care by reducing travel mileage necessary for in-person encounters(23).

Between groups, there were no significant differences in telemedicine cancellation proportions (p > 0.05). This observation indicates that while patients living farther from the clinic rely on telemedicine more frequently, their likelihood of canceling these visits is similar to that of patients living closer. This reflects that telemedicine has the potential to offer a viable means of maintaining access to care for rural patients without increasing appointment non-completion rates.

Interestingly, in-person cancellations were highest among the Short commute group and lowest among Medium and Long commute groups. This pattern may reflect differing levels of commitment or scheduling flexibility, showing that patients who live nearby might be more likely to cancel or reschedule appointments on short notice, whereas those traveling longer distances may plan more intentionally and are thus more likely to attend. In contrast, telemedicine cancellations were consistent across commute groups, suggesting that telemedicine maintains a more stable adherence regardless of travel distance.

Additionally, within each commute group, telemedicine cancellation proportions were less than in-person appointments. Although this is supportive that telemedicine may modestly reduce appointment cancellation it does not take into consideration appointments that must occur in-person and that cannot be done via a telemedicine visit. Taken together, these findings highlight how telemedicine may help standardize access and adherence across geographic contexts, even as behavioral patterns differ among in-person visits.

### Strengths & Limitations

While this study provides valuable insights, it is important to acknowledge its limitations in order to enhance the quality of future research. First, this study only collected data from a single rural family medicine clinic located in the Pacific Northwest of the United States which limits its application outside of this specific demographic. While the 78.5% of patients who identified as White (Table 1) is representative of the U.S. rural population as a whole with 76% White residents, the rest of the patient population is not generally representative, particularly due to the high percentage of patients that answered “Other Race” or declined to answer(24). Although it is possible that race does not significantly affect rural patient adherence to appointment times, further research with a more representative patient population to the US rural population at large would be beneficial.

Second, this study only considered commute times when analyzing patient adherence to appointments. It did not take into account variables such as household income, insurance status, employment status, access to childcare, nor other social determinants of health that could be confounding influences on the proportions of appointment cancellation and Telehealth utilization. As a result, the findings reflect unadjusted associations rather than causal relationships. Future research should incorporate these additional variables and employ study designs that can better control for confounding and bias, such as multivariable regression, propensity matching, or prospective analyses. While these variables should be considered in future research, this study opted to isolate rural patient commute time as this seemed likely to be the strongest predictor of patient adherence to appointment times.

Third, visit type (telemedicine vs. in-person) was determined by patient and provider preference, introducing possible selection bias. Patients who chose telemedicine may differ systematically from those who chose in-person visits in ways that were not measurable in our data set (e.g., health literacy, comfort with technology, or severity of illness).

Despite the limitations on generalizability and conclusivity, a major strength of the single clinic perspective is the unique and intimate ability to gather data on each individual patient rather than general households. This meant the study was able to include and statistically analyze the full range of primary care appointments including pediatric, obstetric, gynecological, geriatric, annual visits, etc. This comprehensive dataset provides a nuanced view of appointment adherence patterns in a rural setting and serves as an important foundation for future multi-site investigations. In doing so, the results of the data are believed to be applicable to similar primary care appointment types in rural areas.

## Conclusion

Examining the relationship between rural patient commute times and appointment adherence was the primary aim of this study. Supported by literature, this study also investigated telemedicine utilization as a potential means of addressing barriers related to long travel times. Long commute times (>30 minutes) were observed to significantly increase the proportion of appointment cancellation compared to medium (15-30 minutes) and short (<15 minutes) commutes, which aligned with the study’s original hypothesis. Long commute patients also utilized telemedicine appointments significantly more than Medium or Short commute patients. As hypothesized, this indicates a strong relationship between travel time and telemedicine adoption. Finally, the proportion of telemedicine appointment cancellations did not differ significantly amongst the different commute time groups, despite the differences in telemedicine utilization proportions. These findings suggest that rural patients who live farther from health care sites increasingly rely on telemedicine to maintain access, without experiencing higher rates of appointment noncompletion. The results of this study add to the current literature addressing the major health disparities rural patients face today. The findings also expound on the current research regarding the potential utilization of telemedicine, which is becoming increasingly more useful and widespread following the COVID-19 pandemic. Future research should seek to continue these efforts within a rural population that is more closely aligned with the demographics of the U.S. rural patient population as a whole.

## Data Availability

The authors do not have permission to distribute the data used as it would risk the privacy of the patients involved. If there are any inquiries about the data, please contact the corresponding author to answer any questions.

## References

1. Nasser HE. One in five Americans live in rural areas. Census.gov. 2017 Aug 9 [cited 2025 Aug 9]. Available from: https://www.census.gov/library/stories/2017/08/rural-america.html

2. Geyman JP, Hart LG, Norris TE, Coombs JB, Lishner DM. Educating generalist physicians for rural practice: how are we doing? J Rural Health. 2000;16(1):56–80. doi:10.1111/j.1748-0361.2000.tb00436.x

3. Cohen S, Metcalf E, Brown MJ, Ahmed NH, Nash C, Greaney ML. A closer examination of the “rural mortality penalty”: variability by race, region, and measurement. J Rural Health. 2025;41(1):e12876. doi:10.1111/jrh.12876

4. Harrington KA, Cameron NA, Culler K, Grobman WA, Khan SS. Rural-urban disparities in adverse maternal outcomes in the United States, 2016-2019. Am J Public Health. 2023;113(2):224–227. doi:10.2105/AJPH.2022.307134

5. Ehrenthal DB, Kuo HD, Kirby RS. Infant mortality in rural and nonrural counties in the United States. Pediatrics. 2020;146(5):e20200464. doi:10.1542/peds.2020-0464

6. Zahnd WE, Fogleman AJ, Jenkins WD. Rural-urban disparities in stage of diagnosis among cancers with preventive opportunities. Am J Prev Med. 2018;54(5):688–698. doi:10.1016/j.amepre.2018.01.021

7. Semprini J, Gadag K, Williams G, Muldrow A, Zahnd WE. Rural-urban cancer incidence and trends in the United States, 2000 to 2019. Cancer Epidemiol Biomarkers Prev. 2024;33(8):1012–1022. doi:10.1158/1055-9965.EPI-24-0072

8. Prissel CM, Grossardt BR, Finney Rutten LJ, Patten CA, Austin JD, St Sauver JL. Midwest urban-rural differences in cancer prevalence. Mayo Clin Proc. 2025;100(7):1172–1187. doi:10.1016/j.mayocp.2024.11.026

9. Wallace DJ, Ray KN, Degan A, Kurland K, Angus DC, Malinow A. Transportation characteristics associated with non-arrivals to paediatric clinic appointments: a retrospective analysis of 51 580 scheduled visits. BMJ Qual Saf. 2018;27(6):437–444. doi:10.1136/bmjqs-2017-007168

10. Maldonado LY, Fryer KE, Tucker CM, Stuebe AM. The association between travel time and prenatal care attendance. Am J Perinatol. 2020;37(11):1146–1154. doi:10.1055/s-0039-1692455

11. Nuako A, Liu J, Pham G, et al. Quantifying rural disparity in healthcare utilization in the United States: analysis of a large midwestern healthcare system. PLoS One. 2022;17(2):e0263718. doi:10.1371/journal.pone.0263718

12. Kurz D, Befort C. Travel burden in a rural primary care behavioral weight loss randomized trial: impact on visit attendance and weight loss. J Rural Health. 2022;38(4):980–985. doi:10.1111/jrh.12652

13. Baldomero AK, Kunisaki KM, Wendt CH, et al. Drive time and receipt of guideline-recommended screening, diagnosis, and treatment. JAMA Netw Open. 2022;5(11):e2240290. doi:10.1001/jamanetworkopen.2022.40290

14. Haggerty T, Stephens HM, Peckens SA, et al. Telemedicine versus in-person primary care: impact on visit completion rate in a rural Appalachian population. J Am Board Fam Med. 2022;35(3):475–484. doi:10.3122/jabfm.2022.03.210518

15. Butzner M, Cuffee Y. Telehealth interventions and outcomes across rural communities in the United States: narrative review. J Med Internet Res. 2021;23(8):e29575. doi:10.2196/29575

16. Ftouni R, AlJardali B, Hamdanieh M, Ftouni L, Salem N. Challenges of telemedicine during the COVID-19 pandemic: a systematic review. BMC Med Inform Decis Mak. 2022;22(1):207. doi:10.1186/s12911-022-01952-0

17. Health Resources and Services Administration. How we define rural [Internet]. U.S. Department of Health and Human Services; 2025 Sep [cited 2026 Feb 2]. Available from: https://www.hrsa.gov/rural-health/about-us/what-is-rural

18. US Department of Health and Human Services. HIPAA privacy rule and research [Internet]. 2002.

19. Al-Sobky RM, Mousa RM. Estimating free flow speed using Google Maps API: accuracy, limitations, and applications. Adv Transp Stud Sect A. 2020;49:39–48. doi:10.4399/97888255317324

20. US Census Bureau. Nation’s urban and rural populations shift following 2020 census [Internet]. 2022 Dec 29 [cited 2025 Aug 9]. Available from: https://www.census.gov/newsroom/press-releases/2022/urban-rural-populations.html

21. Obrochta CA, Parada H Jr, Murphy JD, et al. The impact of patient travel time on disparities in treatment for early stage lung cancer in California. PLoS One. 2022;17(10):e0272076. doi:10.1371/journal.pone.0272076

22. Adepoju OE, Chae M, Ojinnaka CO, Shetty S, Angelocci T. Utilization gaps during the COVID-19 pandemic: racial and ethnic disparities in telemedicine uptake in federally qualified health center clinics. J Gen Intern Med. 2022;37(5):1191–1197. doi:10.1007/s11606-021-07304-4

23. Hahn Z, Hotchkiss J, Atwood C, et al. Travel burden as a measure of healthcare access and the impact of telehealth within the Veterans Health Administration. J Gen Intern Med. 2023;38(Suppl 3):805–813. doi:10.1007/s11606-023-08125-3

24. Johnson K, Lichter D. Growing racial diversity in rural America: results from the 2020 census [Internet]. Carsey School of Public Policy; 2022 May 17 [cited 2025 Aug 9]. Available from: https://carsey.unh.edu/publication/growing-racial-diversity-rural-america-results-2020-census

